# Psychometric Properties of the Spanish Simultaneous Alcohol and Cannabis Motives Measure (S-SACM) among Young Adults

**DOI:** 10.1101/2024.07.16.24310436

**Authors:** Lucía Vélez Pérez, José Carmona Márquez, Bella M. González Ponce, Fermín Fernández Calderón

## Abstract

**Background:** Simultaneous Alcohol and Cannabis (SAC) use is associated with greater negative consequences than individual use. Motives to use are robust predictors of alcohol and cannabis consumption, both separately and simultaneously. While instruments such as the SAC Motives Scale and its short form have recently been developed, no Spanish adaptation is currently available.

**Aims:** To provide a Spanish adaptation of both versions of the simultaneous alcohol and cannabis motives measure and to examine its psychometric properties in a sample of young adults who simultaneously consume both substances.

**Method:** A targeted sampling procedure was used to recruit 612 young cannabis users, of whom 479 reported SAC use (18-25 years old [*M* = 21.01, *SD =* 2.14]; 36% female). Following adaptation and translation, participants completed the scale, along with measures of alcohol, cannabis, and simultaneous use, cannabis motives, and alcohol- and cannabis-related negative consequences.

**Results:** Confirmatory factor analyses showed a four-factor structure (conformity, positive effects, calm/coping, and social). Moreover, the results indicated acceptable internal consistency (.70 - .88), providing evidence of convergent, discriminant, and predictive validity for most dimensions.

**Conclusions:** The results suggest that both versions of the scale are valuable tools for assessing motives for SAC consumption among young adults. These scales could aid in investigating motives for SAC use and informing interventions to mitigate associated harms.

**Public Significance Statement:** This study provides the first Spanish adapted-version measure to assess motives for Simultaneous Alcohol and Cannabis (SAC) consumption. Our findings support the use of this measure for assessing these motives and guiding interventions aimed at preventing or reducing SAC use and its negative consequences among Spanish-speaking young adults who use alcohol and cannabis simultaneously.

---

Alcohol and cannabis rank among the most commonly used psychoactive substances worldwide (World Health Organization [WHO], 2022), particularly in young adults. In Europe, 78.9% of individuals aged 15-24 years reported having consumed alcohol in the past year, while 19.1% reported cannabis use (European Monitoring Centre for Drugs and Drug Addiction [EMCDDA], 2023). A significant percentage of people who use alcohol and cannabis report engaging in joint consumption (i.e., polysubstance use; Brière et al., 2011; Subbaraman & Kerr, 2015), either simultaneously (where effects overlap, operationalized as Simultaneous Alcohol and Cannabis use - SAC) or concurrently (CAC). SAC use, compared to non-simultaneous use, has been linked to greater subjective intoxication and increased negative outcomes such as poor academic performance, absenteeism, committing criminal acts under the influence, impaired relationships, and physical or mental health issues (e.g., symptoms of depression, dependence; Brière et al., 2011; Fleming et al., 2021; Sokolovsky et al., 2020). Moreover, SAC use increases the likelihood of engaging in risky behaviors, including unprotected sex and substance-involved driving (Lee et al., 2022).

According to the motivational model of substance use (Cooper et al., 2016; Cox & Klinger, 1988), motives to consume substances consistently emerge as strong immediate predictors of consumption behavior and its negative consequences, evident in both alcohol (e.g., Patrick & Terry-McElrath, 2021) and cannabis (e.g., Bresin & Mewaki, 2019) use. In the alcohol domain, Cooper (1994) developed the Drinking Motives Questionnaire (DMQ), which identifies four types of drinking motives: social motives (to enhance/facilitate social relationships), enhancement motives (to increase positive affective states), coping motives (to manage negative affective states), and conformity motives (to integrate into a social group). Considering these motives, Simons et al. (1998) devised the Marijuana Motives Measure (MMM), which includes the motives mentioned above along with expansion motives (to heighten awareness/creativity). These two instruments have served as a basis for developing interventions to reduce consumption and its consequences (Banes et al., 2014; Blevins et al., 2016; Canale et al., 2015; Wurdack et al., 2016). Given the mounting evidence for the potential negative consequences of SAC use (Lee et al., 2022), research in this field has analyzed the role of motives for consuming both substances simultaneously. Initially, some studies explored these motives using open-ended questions (e.g., Terry-McElrath et al., 2013), while others have separately assessed alcohol and cannabis use motives (Shipley & Braitman, 2024). To our knowledge, only one standardized instrument is currently available to measure motives for SAC use, the Motives for Simultaneous Alcohol and Marijuana Use Measure, developed by Patrick et al. (2018) in a community sample of young adults in the US.

Patrick et al. (2018) began with an initial pool of 26 items derived from measures of alcohol use motives (Grant et al., 2007), cannabis use motives (Lee et al., 2009), and insights from a review of the perceived effects of SAC use as well as qualitative data collected by their team. Exploratory factor analysis was conducted with a community sample of 286 young adults in the US, resulting in a final 22-item version with a four-correlated factor structure. Three factors (social, calm/coping, and conformity motives) were analogous to those identified for alcohol and cannabis, while a fourth dimension, Positive effects, emerged. This dimension pertains to the use of marijuana to enhance effects when combined with alcohol, aiming to induce a creative or altered state of consciousness (e.g., “Cross-faded effects are better”). This concept draws from alcohol enhancement motives (Cooper, 1994) and marijuana use motives for altered perception (Lee et al., 2009), akin to the expansion motives described in the MMM-SF (Mezquita et al., 2019). Patrick et al. (2018) found that the scale dimensions exhibited stronger associations with analogous dimensions of alcohol and cannabis motives (convergent validity) and weaker associations with other dimensions (discriminant validity).

Two years later, Conway et al. (2020) supported the psychometric soundness of the measure devised by Patrick et al. (2018) and developed a brief version of this scale based on a substantial sample of college students (*n* = 1014) who reported SAC use at least once in the preceding three months. For the brief version, the authors utilized the initial 26-item pool of Patrick et al. (2018). They conducted exploratory and confirmatory factor analyses, providing a brief 11-item version with the same original four-factor structure, the Brief SAM Motives Measure (B-SMM).

In both the full-length (Patrick et al., 2018) and brief (Conway et al., 2020) versions of the scale, social, calm/coping, and positive-effects motives were associated with SAC use. In contrast, conformity motives were linked to decreased SAC use. Additionally, Patrick et al. (2018) observed that coping was the only motive predicting heightened negative consequences of cannabis use (but not alcohol use). Conversely, Conway et al. (2020) reported contrasting findings; calm/coping motives were not correlated with increased SAC-related negative consequences, while the remaining three motives were linked to elevated SAC-related consequences. The subscales of both versions demonstrated satisfactory reliability coefficients (Alpha values between 0.77 and 0.88).

Given the unavailability of both the full-length and short-form versions of the Motives for Simultaneous Alcohol and Marijuana Use Measure in Spanish, our objective was to provide a Spanish adaptation of the SAC motives measure (Patrick et al., 2018) and its short-form (Conway et al., 2020). Subsequently, we sought to assess its psychometric properties within a community-based sample of young adults who reported SAC use in the past month. Specifically, our aims included establishing evidence of reliability (internal consistency) and validity in terms of its internal structure and its correlations with other variables, such as motives for cannabis use, SAC use, and alcohol and cannabis use and related consequences.

## Method

### Participants and procedure

We employed a targeted sampling method (Watters & Bernacki, 1989) to recruit a community-based sample of 616 young adults in Seville and Huelva, Spain, who reported cannabis use within the past month. The final sample comprised 612 participants (aged 18-25 [*M* = 21.04, *SD* = 2.16]), with 60.6% identifying as male, following the exclusion of four individuals displaying inconsistent response patterns. Initially, potential sampling locations frequented by the target population were identified across Huelva and Seville, such as parks, plazas, and bars/pubs. For this purpose, young adults with similar characteristics to the target population were interviewed, and a list of these contexts was created and categorized by districts across both cities to favour socioeconomic diversity of the participants.

During the subsequent phase, three psychologists with research expertise in social psychology visited the designated sites and walked the streets within the identified districts. They approached individuals who appeared to meet the age criterion (i.e., 18-25 years) and invited them to participate. Subsequently, those who expressed interest and met this age criterion were contacted via telephone to ascertain if they met a second inclusion criterion (i.e., having consumed cannabis at least once in the last month). Additionally, posters containing basic information about the study and a contact telephone number were prominently displayed in the selected settings.

Within the targeted sampling framework, snowball sampling (Goodman, 1961) was used for sample recruitment, with each participant restricted to nominating up to five individuals to avoid sample homogeneity. Additionally, an adaptive sampling approach (Thompson & Collins, 2002) was adopted, whereby certain participant characteristics were assessed parallel to the selection process to ensure a diverse sample in terms of sex, age, university status, and cannabis use patterns. Of the total participant pool (n = 612), 286 (46.7%) responded to the posters by contacting the interviewer, 198 (32.4%) were referred by other participants, and 128 (21.0%) were recruited in person by the interviewer.

Participants were required to provide written consent before completing a self-administered questionnaire in the presence of the interviewer, individually or in small groups (five people or fewer). Data were collected in rooms at the University of Huelva and collaborating organizations specializing in substance use in Seville. As a token of appreciation, each participant received a 15-euro Amazon voucher in printed format. The study protocol was approved by the Regional Bioethics Research Committee of Andalusia (Consejería de Sanidad, Junta de Andalucía, Spain).

For the current study, the analytical sample comprised 479 individuals (78.3% of the total sample) who reported SAC use within the last month (Mean age = 21.01, *SD* = 2.14; 36.6% female). Among these participants, 42.0% reported being enrolled in university at the time of the study, and the majority (64.5%) resided with their parents, with family being their main source of financial support (48.2%). On average, participants reported SAC consumption on 5.34 days within the last month (*SD* = 5.51).

### Instruments

#### Spanish version of the Simultaneous Alcohol and Cannabis Motives Measure

The adaptation process adhered to the guidelines outlined by the International Test Commission (ITC, 2017), considering the cultural, psychological, and linguistic differences between the United States and Spain. It is worth noting that in Spain, both marijuana and hashish are frequently used (Observatorio Español de las Drogas y las Adicciones [OEDA], 2023). Consequently, the decision was made to replace the term “marijuana” with “cannabis” in both the instructions and wording of the items to ensure cultural relevance and linguistic appropriateness for the Spanish context.

The process of translating and adapting the instrument took place across four phases, overseen by three research team members with expertise in psychometrics and substance use research. Initially, the corresponding author drafted a document containing the 22 original items alongside three blank columns for each researcher to independently propose initial translations for each item. Subsequently, in the second phase, the corresponding author collated these initial proposals with the original items and circulated them among the remaining researchers, who contributed a second version, considering the team’s suggestions. Upon achieving consensus on 18 of the 22 items, the third phase entailed a face-to-face meeting among team members to discuss the remaining four items until consensus was reached. This version was then back-translated by an American professional translator (PhD in Psychology) in the fourth phase, resulting in slight modifications to the wording of some items. Consistent with Patrick et al. (2018) and Conway et al. (2020), a five-point Likert-type response format (1 = almost never/never, 5 = almost always/always) was employed. See Supplemental Appendix-A for the complete versions of the S-SACM and S-SACM-SF, including application instructions.

#### Frequency of Alcohol, Cannabis, and Simultaneous Alcohol and Cannabis Use

Participants were asked about the number of days on which they had used cannabis and alcohol simultaneously (“on the same occasion so that the effects overlap”) in the past 30 days. In addition, the number of days of cannabis use and alcohol use in the past month were measured independently.

#### Quantity of Alcohol Used in a Typical Week during the Past Month

To assess the quantity of alcohol intake over a typical week, a modified version of the Daily Drinking Questionnaire (DDQ, Collins et al., 1985) was administered. Following the guidelines established by the Spanish Observatory on Drug and Addiction (OEDA, 2019), participants were asked about their consumption of six varieties of alcoholic beverages, accompanied by images of each beverage. Subsequently, the quantity consumed for each beverage type was converted into its corresponding Standard Drinking Units (SDU), with each unit in Spain equating to 10 grams of pure alcohol (Llopis-Llácer et al., 2000).

#### Quantity of Cannabis Used in a Typical Week during the Past Month

The Marijuana Use Grid (MUG, Pearson & Marijuana Outcomes Study Team, unpublished) was administered, featuring a table with each day of the week divided into six four-hour intervals. Participants were required to indicate the grams of cannabis consumed during each period, separately for hashish and marijuana. The grams reported for each interval and day were summed to compute the total amount of cannabis consumed over a typical week. To aid participants in estimating the grams consumed, an image was provided displaying various quantities of hashish and marijuana (0.25g / 0.5g / 1.0g / 2.5 g) next to a soda cap (refer to Figure A1 in Supplemental Appendix A).

#### Cannabis Use Motives

We employed the Marijuana Motives Measures Short-Form (MMM-SF; Simons et al., 1998), adapted to Spanish by Mezquita et al. (2019). This scale comprises 15 items rated on a five-point scale (1 = almost never or never, 5 = almost always or always), organized into five dimensions (each consisting of three items): social motives, coping motives, enhancement motives, conformity motives, and expansion motives. The Cronbach’s alpha values for each dimension were as follows: social motives (α = .77), coping motives (α = .77), enhancement motives (α = .56), conformity motives (α = .76), and expansion motives (α = .84).

#### Cannabis-related Negative Consequences

The Brief Marijuana Consequences Questionnaire (B-MACQ; Simons et al., 2012), in its Spanish version (Bravo et al., 2019), was employed to assess cannabis-related negative consequences experienced in the last month. The B-MACQ comprises 21 dichotomous items (presence = 1 / absence = 0) that evaluate various negative consequences, with scores summed to derive a total score. As recommended by Bravo et al. (2019), Item 5 (“I have gotten into physical fights because of my marijuana use”) was excluded. The Cronbach’s alpha for our sample was .80.

#### Alcohol-related Negative Consequences

We used the Spanish version (Pilatti et al., 2014) of the Brief Young Adults Alcohol Consequences Questionnaire (B-YAACQ; Kahler et al., 2005), which consists of 24 dichotomous items (presence = 1 / absence = 0). Responses were summed to obtain a total score of alcohol-related problems. The Cronbach’s alpha for our sample was .85.

### Data Analysis

Confirmatory Factor Analyses (CFAs) were conducted to evaluate the fit of the scale in its full version (22 items) to the four correlated factor structure proposed by Patrick et al. (2018). Additionally, the fit of this model was compared to that of several alternative models, including a one-factor model, a four-factor uncorrelated model, and a model comprising one second-order factor and four first-order factors. The fit of the items in the brief version (11 items, Conway et al., 2020) to the four-factor correlated structure and the same alternative models was also assessed.

The CFAs were conducted in Mplus 8.7 (Muthén & Muthén, 1998-2017) using the weighted least squares mean and variance adjusted (WLSMV) estimation method. To assess the overall fit of the models, the robust chi-square, the Comparative Fit Index (CFI), the Tucker-Lewis Index (TLI), the Root Mean Square Error of Approximation (RMSEA), and the Standardized Root Mean Square Residual (SRMR) were used. CFI and TLI values greater than .95, RMSEA values lower than .06, and SRMR values lower than .08 indicate a good fit (Hu & Bentler, 1999). Specific sources of model misspecification were initially identified based on Modification Indices (MI) and Standardized Estimated Parameter Changes (EPC). Additionally, an Exploratory Structural Equation Modeling (ESEM) approach was employed. Items with standardized loadings lower than |0.4| on their targeted factor or standardized cross-loadings greater than |0.3| on non-targeted factors in the ESEM were further examined (Steenkamp & Maydeu-Olivares, 2023). The DIFFTEST procedure in Mplus was used to compare the fit of nested models.

To provide evidence of the validity of both the full-length and short-form versions based on their relationships with other variables, Pearson’s correlations were employed. These correlations examined the associations between dimensions of SAC motives and several variables, including SAC use (number of days of SAC use in the past month), cannabis motives (social, enhancement, coping, conformity, and expansion), cannabis-related outcomes (frequency of cannabis use, quantity of cannabis consumed, and cannabis-related negative consequences), and alcohol-related outcomes (frequency of alcohol use, quantity of alcohol consumed, and alcohol-related negative consequences). Cronbach’s alpha reliability for each subscale score was estimated using SPSS 28.0 (IBM Corp., 2021), along with the conduct of correlation analyses.

### Transparency and Openness

The design and analysis of this study were not preregistered. All data, materials and analysis code have been made publicly available at the Open Science Framework and can be accessed at [https://osf.io/u79pr/?view_only=b60b32832def4395bc482f365d387a2b].

## Results

### Item Analysis

As displayed in Table 1, the S-SACM items showed corrected item-test correlation values ranging from .24 to .79. In the S-SACM-SF, these values ranged from .40 to .77.

**Table 1.** Mean, standard deviation and item-total correlation of the items of both versions of the Spanish SAC Motives Measure.

| Subscales/Items | *M* | *SD* | Corrected Item-total correlation |  |
| --- | --- | --- | --- | --- |
| Conformity |  |  | 22-item | 11-item |
| 1. Because others are doing it [Porque otras personas lo están haciendo] | 1.94 | 1.20 | .29 |  |
| 2. Pressure from others [Por la presión de otras personas] | 1.24 | 0.61 | .72 | .68 |
| 3. Looking for a new experience [Por buscar una nueva experiencia] | 1.92 | 1.20 | .24 |  |
| 4. So I won't feel left out [Para no sentirme excluido/a] | 1.21 | 0.62 | .69 |  |
| 5. To fit in with a group I like [Para encajar en un grupo que me gusta] | 1.15 | 0.51 | .66 | .72 |
| 6. Because my friends pressure me to use [Porque mis amigos/as me presionan para que lo haga] | 1.15 | 0.52 | .60 |  |
| 7. So that others won't kid me about not using [Para que los demás no se burlen de mi por no hacerlo] | 1.07 | 0.34 | .56 | .66 |
| 8. To be liked [Para gustar a los demás] | 1.10 | 0.46 | .59 |  |
| Sum of scores (8 – 36) | 10.79 | 3.59 |  |  |
| Sum of scores (3 – 14) | 3.46 | 1.30 |  |  |
| Positive Effects |  |  |  |  |
| 9. To increase intoxication [Para colocarme más] | 3.44 | 1.39 | .66 |  |
| 10. Cross-faded effects are better [Porque los efectos de mezclarlas son mejores] | 2.94 | 1.30 | .72 | .59 |
| 11. To get a better high [Para tener un mejor colocón] | 3.20 | 1.33 | .79 |  |
| 12. To get to a greater altered state [Para alterar más mi consciencia] | 2.54 | 1.36 | .57 |  |
| 13. To increase effects I get from alcohol [Para aumentar los efectos positivos del alcohol] | 2.51 | 1.38 | .69 | .77 |
| 14. To increase effects I get from cannabis [Para aumentar los efectos positivos del cannabis] | 2.70 | 1.42 | .69 | .74 |
| Sum of scores (6 - 30) | 17.32 | 6.41 |  |  |
| Sum of scores (3 – 15) | 8.14 | 3.55 |  |  |
| **Calm/Coping** |  |  | 22-item | 11-item |
| 15. To help me sleep [Porque me ayuda a dormir] | 2.10 | 1.40 | .67 |  |
| **16. To calm down [Para calmarme]** | 1.93 | 1.24 | .79 | .70 |
| **17. To cope with anxiety [Para afrontar la ansiedad]** | 1.74 | 1.18 | .67 | .70 |
| Sum of scores (3 - 15) | 5.77 | 3.33 |  |  |
| Sum of scores (2 – 10) | 3.67 | 2.23 |  |  |
| **Social** |  |  |  |  |
| **18. Because it makes a social gathering more enjoyable [Porque hace que las reuniones sociales sean más divertidas]** | 3.20 | 1.39 | .48 | .42 |
| **19. Because it is customary on special occasions [Porque es lo que se suele hacer en ocasiones especiales]** | 2.53 | 1.43 | .61 | .57 |
| 20. To be sociable [Para ser más sociable] | 1.99 | 1.21 | .44 |  |
| 21. Because it is what most of my friends do when we get together [Porque es lo que la mayoría de mis amigos/as hace cuando nos reunimos] | 2.05 | 1.22 | .36 |  |
| **22. As a way to celebrate [Para celebrar algo]** | 3.21 | 1.44 | .38 | .40 |
| Sum of scores (5 - 24) | 12.97 | 4.51 |  |  |
| Sum of scores (3 – 15) | 8.94 | 3.26 |  |  |

*Note.* Items included in the short version are shown in bold.

### Evidence of validity based on the internal structure and reliability of scale scores

The four-factor correlated model showed an adequate fit to the 22 scale items, yielding WLSMV *χ^2^* (203) = 720.01, *p* < .01, CFI = .95, TLI = .95, RMSEA = .07 and SRMR = .08 (Table 2). Moreover, this model showed a significantly superior fit compared to the other models evaluated.

**Table 2.** Goodness-of-Fit Indices of the CFA models for the full-length version of the Spanish SAC Motives Measure (S-SACM) and the Spanish SAC Motives Measure Short-Form (S-SACM-SF)

| Models | WLSMV $\chi^2$ (df) | CFI | TLI | RMSEA [90% CI] | SRMR | DIFFTEST $\chi^2$ (df) |
| --- | --- | --- | --- | --- | --- | --- |
| S-SACM |  |  |  |  |  |  |
| Four Correlated Factors | 720.01* (203) | .95 | .95 | .07 [.07, .08] | .08 |  |
| One Factor | 2719.17* (209) | .77 | .75 | .16 [.15, .16] | .23 | 575.36* (6) |
| Four Uncorrelated Factors | 1506.37* (209) | .88 | .87 | .11 [.11, .12] | .16 | 245.68* (6) |
| Second Order | 717.10* (205) | .95 | .95 | .07 [.07, .08] | .09 | 18.72* (2) |
| S-SACM-SF |  |  |  |  |  |  |
| Four Correlated Factors | 88.50* (38) | .99 | .99 | .05 [.04, .07] | .04 |  |
| One Factor | 1072.28* (44) | .80 | .75 | .22 [.21, .23] | .21 | 469.10* (6) |
| Four Uncorrelated Factors<sup>1</sup> | 446.65* (45) | .92 | .90 | .14 [.13, .15] | .13 | 188.12* (7) |
| Second Order | 127.01* (40) | .98 | .98 | .07 [.05, .08] | .07 | 20.22* (2) |

*Note.* Estimation Method: Weighted Least Square Mean and Variance Adjusted (WLSMV); CFI = Comparative Fit Index; TLI = Tucker Lewis Index; RMSEA = Root Mean Square Error Approximation; SRMR = Standardized Root Mean Square Residual.
<sup>1</sup> The loadings of the two items of the Calm/Coping Factor were constrained to be equal for identification purposes.
\* $p < .05$ ; \*\* $p < .01$ .

All factor loadings in the long version were statistically significant and greater than .40 (Figure 1). The correlations between factors ranged between .15 and .53 and were all statistically significant.

**Figure 1.**
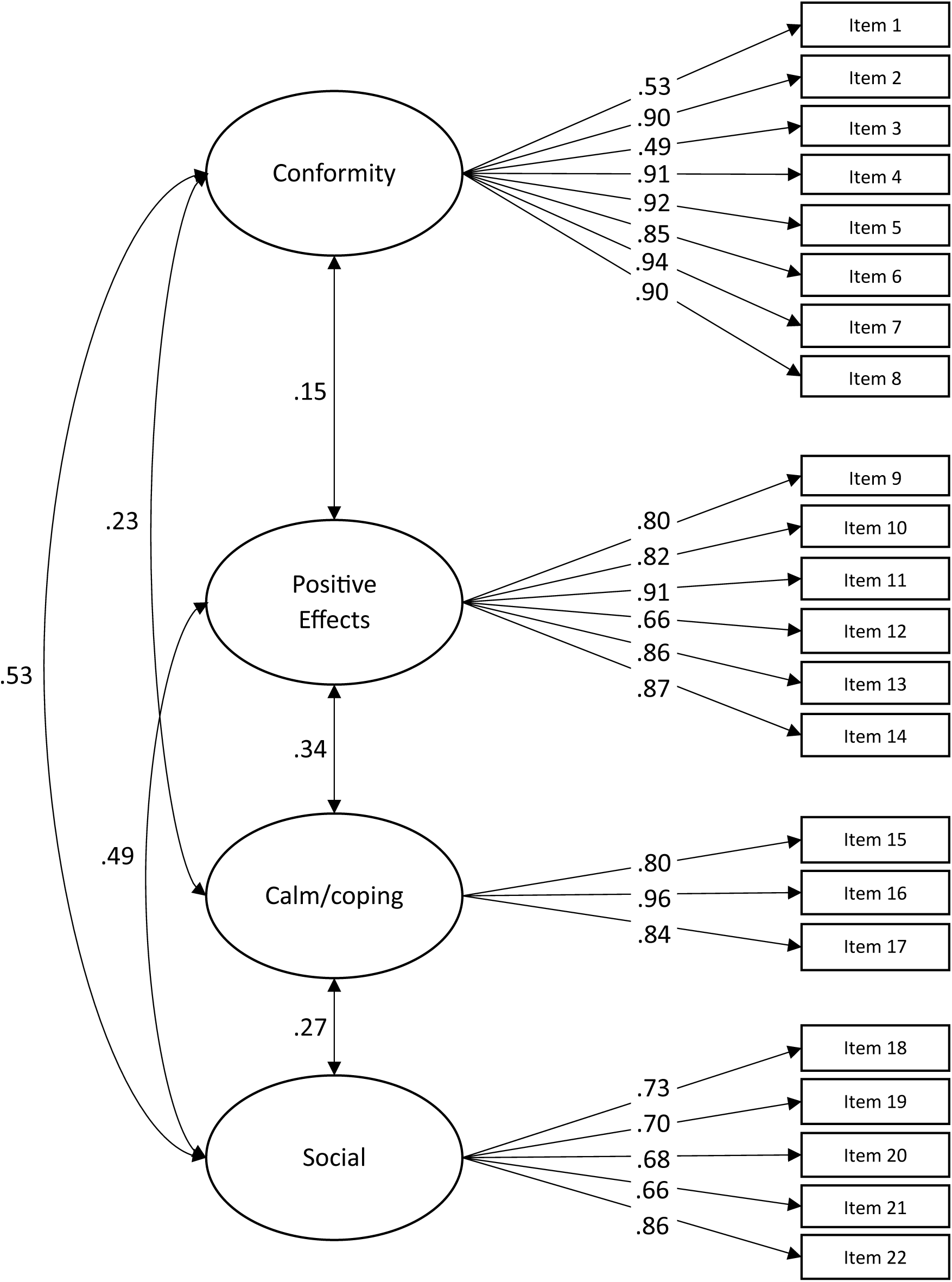
Standardized parameters of the four factors CFA model for the 22 items of the Spanish SAC Motives Measure (S-SACM)

Next, potential sources of model specification error were explored. The analysis of the *MIs* and *EPCs* revealed that including certain cross-loadings for items 3, 18, and 21, as well as the correlation between the errors of items 13 and 14, could improve the model fit. These potential sources of misspecification were partially confirmed through the analysis of the ESEM parameters. Specifically, item 3 showed a factor loading of .39 (< .40) on its targeted factor (Conformity) and .35 (> .30) on the non-targeted factor of Positive Effects. Item 21 also showed a factor loading of .35 on the non-targeted Conformity factor.

The four correlated factor model for the 11-item short-form version showed a good fit (WLSMV *χ^2^* (38) = 88.50, *p* < .01, CFI = .99, TLI = .99, RMSEA = .05 and SRMR = .04) and outperformed the alternative models (see Table 2). The factor loadings were all significant and, except for Item 22, were all greater than .70 (Figure 2). The analysis of MIs, EPCs, and ESEM parameters revealed no significant local misspecification.

**Figure 2.**
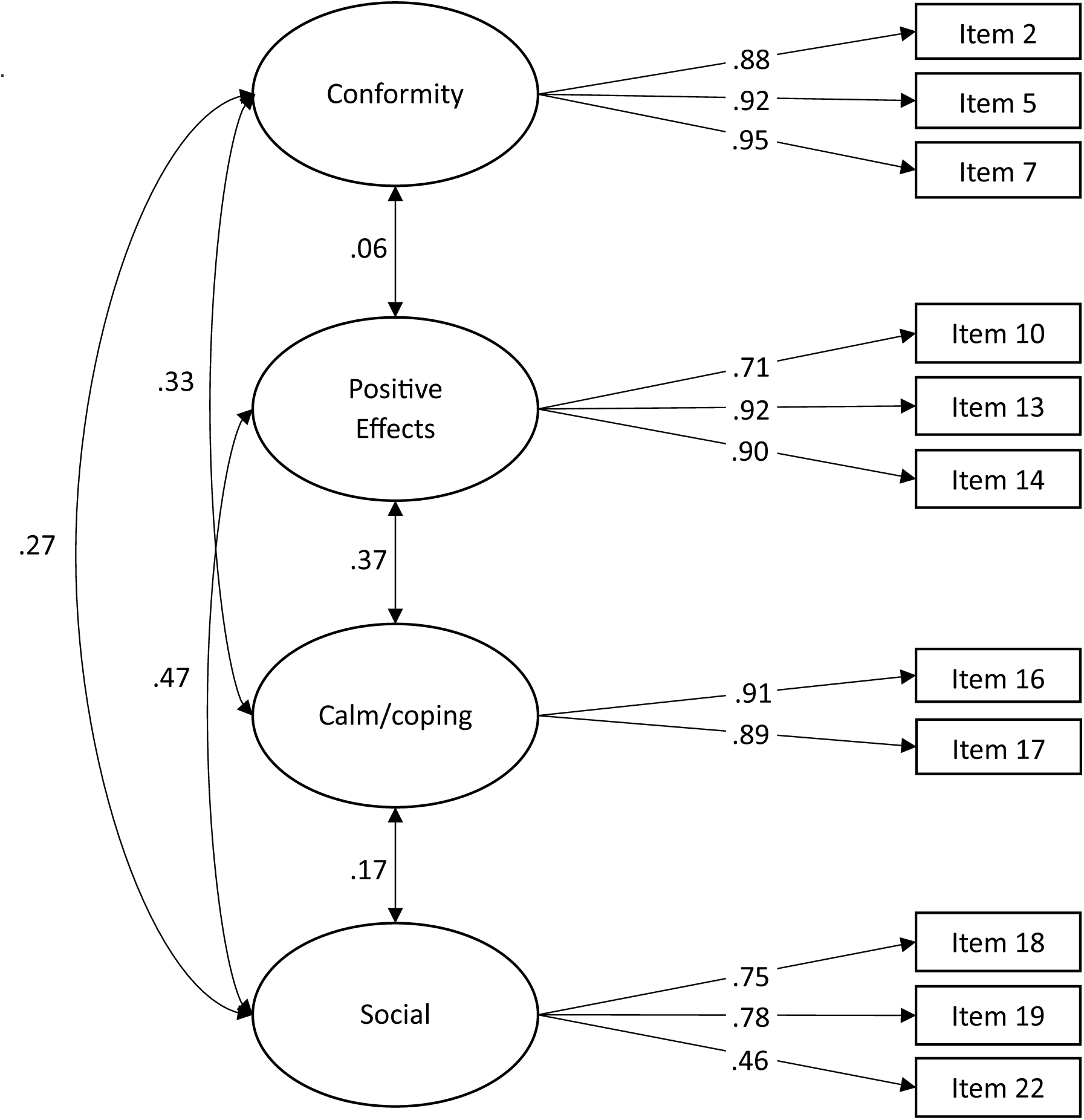
Standardized parameters of the four factors CFA model for the 11 items of the Spanish Short Form SAC Motives Measure (S-SACM-SF)

Internal consistency analyses revealed alpha values for the full-length and short-form versions of .74/.81 (conformity), .84/.82 (calm/coping), .88/.84 (positive effects), and .70/.65 (social).

### Evidence of validity based on the relationship with other variables

As depicted in Table 3, for both the full-length and short-form versions, the various SAC motives showed stronger correlations with their analogous dimensions on the cannabis motives scale and weaker correlations with the remaining dimensions. In both versions, calm/coping motives showed positive associations with all cannabis outcomes. In contrast, positive effects motives were linked with the frequency of SAC use, cannabis-related consequences, and the frequency and quantity of alcohol use, along with its associated consequences. Additionally, conformity and social motives were associated with alcohol and cannabis-related consequences, with conformity motives being the only dimension not correlated with the frequency of SAC use.

**Table 3.** Relationship between SAC motives and other variables.

| Variables | Spanish SAC Motives Measure (S-SACM) |  |  |  | Spanish SAC Motives Measure Short Form (S-SACM-SF) |  |  |  |
| --- | --- | --- | --- | --- | --- | --- | --- | --- |
|  | Conformity | Positive Effects | Calm/coping | Social | Conformity | Positive Effects | Calm/coping | Social |
| Cannabis motives |  |  |  |  |  |  |  |  |
| Conformity | .42** | .11* | .06 | .23** | .43** | .10* | .09 | .16** |
| Enhancement | .05 | .35** | .03 | .24** | -.01 | .25** | .02 | .24** |
| Expansion | .10* | .15** | .11* | .16** | .03 | .13** | .07 | .15** |
| Coping | .11* | .10* | .35** | .19** | .13* | .10* | .40** | .14** |
| Social | .16** | .30** | .03 | .46** | .07 | .28** | .03 | .43** |
| SAC |  |  |  |  |  |  |  |  |
| Frequency of use<sup>a</sup> | -.03 | .14** | .28** | .11* | -.01 | .14** | .26** | .07 |
| Cannabis |  |  |  |  |  |  |  |  |
| Frequency of use<sup>a</sup> | -.10* | .00 | .18** | .07 | -.01 | -.01 | .16** | .08 |
| Quantity of use | .00 | .02 | .15** | .08 | .06 | .04 | .14** | .09* |
| Related consequences | .15** | .13** | .23** | .17** | .14** | .10* | .28** | .11* |
| Alcohol |  |  |  |  |  |  |  |  |
| Frequency of use<sup>a</sup> | .04 | .11* | .26** | .04 | .05 | .13** | .24** | .01 |
| Quantity of use | .00 | .18** | .23** | .12** | .02 | .19** | .19** | .12** |
| Related consequences | .16** | .19** | .33** | .19** | .11* | .18** | .32** | .14** |

*Note.* $N = 479$ (except for quantity of cannabis: $n = 477$ ).
<sup>a</sup> *Frequency of use* has been operationalized as the number of days of use in the last month.
\* $p < .05$ ; \*\* $p < .01$ .

## Discussion

This study provides the first Spanish-adapted version of the SAC motives measure (Patrick et al., 2018), the S-SACM, and its short form (Conway et al., 2020), the S-SACM-SF, in a community sample of cannabis-using young adults. Both versions showed a good fit to the original four-factor correlated structure, with adequate internal consistency and evidence of validity based on their relationships with other variables.

The CFA results for both versions indicated a good fit to the original four-factor correlated model, significantly better than the four-factor uncorrelated model, the one-factor model, and the model including a second-order factor. However, a more detailed analysis of the structure revealed that the fit could be improved by including a correlation between the errors of items 13 and 14. This observation could be attributed to the similarities in wording and content between these items, particularly in their assessment of SAC use to amplify the effects obtained from cannabis (Item 13) and alcohol (Item 14). Moreover, our findings indicated that including cross-loadings for Items 3 (targeted factor=conformity, non-targeted factor=positive effects) and 21 (targeted factor=social, non-targeted factor=conformity) improved the model fit for the full-length version (these two items were not retained in the short-form version). From a conceptual standpoint, the content of Item 3 (“looking for a new experience”) appears unrelated to the targeted factor (Conformity) and is more closely associated with the untargeted factor, Positive effects. Regarding Item 21 (“because it is what most of my friends do when we get together”), it conceptually seems to match both, the expected factor Social motives, and the unexpected factor Conformity motives. Thus, the low corrected item-total correlations for Items 3 and 21 (.24 and .36, respectively), together with the similar findings reported by Patrick et al. (2018), highlight the need for further examination of these two items within the full-length version of the SAC motives measure.

Similar to Patrick et al. (2018) and Conway et al. (2020), our findings demonstrated acceptable/good internal consistency for the motives subscales in both versions. Additionally, evidence of convergent and discriminant validity was established through positive correlations between motives for SAC use and motives for cannabis use, with stronger correlations observed between analogous dimensions of the two scales compared to the rest of the dimensions. These observations are in line with the findings of Patrick et al. (2018), suggesting that both the S-SACM and the S-SACM-SF capture specific motives for SAC use, consistent with the motivational model of substance use (Cooper, 2016).

In both the S-SACM and the S-SACM-SF, SAC motives have shown evidence of validity based on their expected relationships with other variables. Partially consistent with the psychometric study by Conway et al. (2020), higher scores in positive effects motives were associated with more frequent SAC use. This observation is consistent with relationships found in the alcohol domain for enhancement motives (Bresin & Mewaki, 2020) and in the analogous enhancement and expansion motives for cannabis use (Bresin & Mewaki, 2019). However, contrary to the results reported by Patrick et al. (2018), our findings revealed a positive association between social and positive effects motives and alcohol and cannabis-related consequences.

Concerning calm/coping motives, these emerged as the only SAC motives significantly associated with every outcome examined in our study (i.e., SAC, cannabis and alcohol use, alcohol and cannabis-related consequences). Compared to the other SAC motives, calm/coping motives showed the highest correlations with each outcome. These results are partially similar to those reported by Patrick et al. (2018) and Conway et al. (2020) and are consistent with a growing body of evidence indicating that cannabis-coping motives and alcohol-coping motives are strongly associated with the use of both substances and the related consequences (Bresin & Mewaki, 2019; 2020). However, in contrast to the original results (Patrick et al., 2018; Conway et al., 2020), where lower conformity motives predicted higher SAC use, our study found no relationship between consumption and these motives. Previous literature exploring the role of conformity motives for alcohol use and cannabis use has yielded mixed findings, with some studies showing a positive relationship (Lee et al., 2009; Loose et al., 2017), while others reported a negative (Bresin & Mewaki, 2019; Crutzen & Kuntsche, 2013), or no relationship (Bonar et al., 2017; Bonn-Miller, 2007; Kuntsche et al., 2008). In our study, while conformity motives were positively related to both alcohol and cannabis consequences, they were the only SAC motives not related to the frequency of SAC use. In contrast, the studies by Patrick et al. (2018) and Conway et al. (2020) found that higher conformity motives were associated with lower SAC use. Although our findings do not fully support this pattern of relationships, they do indicate that higher conformity motives are associated with a lower frequency of cannabis use.

### Limitations and future research

These results should be interpreted in the light of certain limitations. First, the absence of a frame of reference for young adult SAC consumers in community contexts prompted us to use a non-probabilistic sampling procedure, limiting the ability to generalize the results to all young Spanish SAC consumers. However, to enhance variability and obtain a sample with characteristics and SAC consumption patterns similar to those of the young Spanish population, we analyzed certain sociodemographic characteristics (sex, age, and university level) and the frequency of SAC consumption in parallel with sample selection. Additionally, the cross-sectional design of this study precludes making predictive inferences from the results. Future research should assess the scale’s predictive validity through studies with prospective data. Furthermore, the lack of a specific measure of motives for alcohol consumption prevented us from exploring the relationship between these and the motives for SAC use in our sample. Thus, future studies should include information on the relationship between motives for alcohol use and SAC use to further support the conceptual validity of the scale. Finally, given the absence of a standardized measure of problems specifically arising from SAC use (Shipley & Braitman, 2024), we assessed these issues through measures of the negative consequences of alcohol and cannabis independently. While this has been the predominant strategy used thus far (Shipley & Braitman, 2024), according to the motivational model (Cooper, 2016), the existence of a scale for measuring the negative consequences of SAC use would enable a more accurate assessment of the relationships between SAC motives and the specific negative consequences associated with this pattern of use. Therefore, future investigations should focus on developing this specific measure to obtain evidence of criterion validity (i.e., the extent to which SAC consumption motives accurately predict the associated negative consequences).

### Conclusions

Our findings provide psychometric evidence supporting the reliability and validity of the Spanish adaptation of the SAC use motives scale, both in its full and brief versions, in a community sample of young adults reporting simultaneous alcohol and cannabis use. These results suggest that the S-SACM and the S-SACM-SF are suitable instruments for assessing SAC use motives in research settings. Given the identified associations, these scales could serve as valuable tools for guiding interventions to reduce SAC use and mitigating its negative consequences (e.g., through interventions such as alternative coping skills training) among young adult SAC users.

## Data Availability

All data, materials and analysis code have been made publicly available at the Open Science Framework.

https://osf.io/u79pr/?view_only=b60b32832def4395bc482f365d387a2b

## Acknowledgments

This work was funded by the Agencia Estatal de Investigación (Ministerio de Ciencia, Innovación y Universidades, Spain; MICIU/AEI//10.13039/501100011033/) under grant number PID2020-118229RB-I00 (Principal Investigator: Fermín Fernández Calderón).

The authors would like to thank Federación Andaluza ENLACE and Asociación Nazarena de Terapia de Apoyo, Rehabilitación e Inserción Social (ANTARIS) for their invaluable support.

## Notes

We have no known conflict of interest to disclosure.

### Competing Interest Statement

The authors have declared no competing interest.

### Funding Statement

This work was funded by the Agencia Estatal de Investigacion (Ministerio de Ciencia, Innovacion y Universidades, Spain; MICIU/AEI//10.13039/501100011033/) under grant number PID2020-118229RB-I00 (Principal Investigator: Fermin Fernandez Calderon).

### Author Declarations

The Regional Bioethics Research Committee of Andalusia (Consejeria de Sanidad, Junta de Andalucia, Spain) gave ethical approval for this work.

